# Prevalence of Asymptomatic non-Falciparum and Falciparum Malaria in the 2014-15 Rwanda Demographic Health Survey

**DOI:** 10.1101/2024.01.09.24301054

**Authors:** Claudia Gaither, Camille Morgan, Rebecca Kirby, Corine Karema, Pierre Gashema, Samuel J. White, Hillary M. Topazian, David Geibrecht, Kyaw Thwai, Neeva Wernsman Young, Koby Boyter, Tharcisse Munyaneza, Claude Mambo Muvunyi, Jean De Dieu Butera, Jeffrey A. Bailey, Jean-Baptiste Mazarati, Jonathan J. Juliano

## Abstract

**Background:** Recent molecular surveillance suggests an unexpectedly high prevalence of non-falciparum malaria in Africa. Malaria control is also challenged by undetected asymptomatic *P. falciparum* malaria resulting in an undetectable reservoir for potential transmission. Context-specific surveillance of asymptomatic *P. falciparum* and non-falciparum species is needed to properly inform malaria control programs.

**Methods:** We performed quantitative real time PCR for four malaria species in 5,050 primarily adult individuals in Rwanda using the 2014-2015 Demographic Health Survey. We assessed correlates of infection by species to explore attributes associated with each species. Asymptomatic *P. ovale spp*., *P. malariae, and P. falciparum* malaria infection had broad spatial distribution across Rwanda. *P. vivax* infection was rare.

**Results:** Overall infection prevalence was 22.3% (95%CI [20.3, 24.3]), with *P. falciparum* and non-falciparum at 16.3% [14.5, 18.1] and 8.0% [6.6, 9.3], respectively. Parasitemias tended to be low and mixed species infections were common, especially where malaria transmission and overall prevalence was the highest. *P. falciparum* infection was associated with lower wealth, rural residence and low elevation. Fewer factors were significantly associated with non-falciparum malaria.

**Conclusions:** Asymptomatic non-falciparum malaria and *P. falciparum* malaria are common and widely distributed across Rwanda in adults. Continued molecular monitoring, preferably done by the national malaria control program, of *Plasmodium* diversity using routine survey samples is needed to strengthen malaria control.

## Introduction

In 2023 there were an estimated 263 million malaria cases worldwide, with 94% occurring in the WHO African Region. [1] Human malaria infection is caused primarily by five species of the *Plasmodium* parasite (falciparum, malariae, ovale curtisi, ovale walkeri, and vivax), with most cases and morbidity in Africa attributable to *Plasmodium falciparum*. The clinical burden of non-falciparum species in Africa is not well characterized, but all five species have been reported in multiple countries where they have previously not been considered a threat. Falciparum clearly remains the primary cause of morbidity and mortality, but severe disease has been reported with non-falciparum species as well, including anemia, renal failure and other severe outcomes.[2–5] Increase in non-falciparum prevalence has been observed in areas where falciparum prevalence has steadily declined, presenting opportunities for other Plasmodium species to flourish. [6,7]

Diagnosis of malaria in Africa relies principally on rapid diagnostic tests (RDTs) detecting the antigen *P. falciparum* histidine rich protein 2 (HRP2), often with a second less sensitive band for pan-species lactate dehydrogenase (LDH).[8] Designed to detect clinical malaria, RDTs for malaria surveillance will often miss low density *P. falciparum* infections and infections with non-falciparum species. Alternate approaches are needed to accurately define the full asymptomatic reservoir. Molecular detection, such as through real-time PCR, can identify infections at lower parasitemia levels than RDTs can, and better characterize non-falciparum malaria infections. Molecular approaches are particularly important for use in asymptomatic community surveillance.[9] Asymptomatic infections may have significant but underestimated morbidity,[10] and are often followed by symptomatic infection.[11] In addition, they provide a reservoir for ongoing transmission.[12–14] Thus, characterizing the epidemiology of asymptomatic malaria is important for malaria control and prevention, as malaria elimination requires control of all species of malaria, whether symptomatic or asymptomatic.

Molecular identification of non-falciparum malaria infections (*P. malariae, P. ovale curtisi, P. ovale wallikeri*, and *P. vivax*) is critical for addressing morbidity from these parasites and for reaching malaria elimination in settings with falciparum-based diagnostics and treatments. Accurate diagnosis is imperative for malaria control, as the clinical features and treatments differ. In contrast with *P. falciparum* infection, *P. vivax* and *P. ovale* cause relapse through the persistence of hypnozoites (dormant liver stage parasites).[15,16] As hypnozoites do not respond to blood-stage treatment, namely artemisinin-combination therapies (ACTs), the primary treatment for severe malaria in most countries, relapsing infections require radical cure.[17] Thus, their presence may require national malaria control programs to alter therapeutic options in the country. To-date, *P. ovale spp*. and *P. malariae* malaria have been more frequently described, due in part to wider implementation of highly sensitive nucleic acid detection, and also likely due to increasing prevalence as *P. falciparum* is controlled as was seen for *P. vivax* in Southeast Asia.[6,7,18] *P. vivax* is also being reported more frequently due to wider implementation of molecular diagnostics.[4,19,20]

Rwanda has historically had strong malaria control, leveraging effective antimalarials, insecticide-treated bed nets, and indoor residual spraying.[21] Malaria cases in Rwanda increased from 48 to 403 per 1,000 between 2012 and 2018, with mortality increasing 41% over the same time.[21] Emerging insecticide resistance, an increase in irrigated agriculture, and insufficient insecticide-treated bed net coverage, among other factors, have been attributed to these increases.[21] From 2018 to 2022, malaria prevalence decreased from an estimated 321 to 76 cases per 1,000, still higher than 2012 estimates,[22] and with detection of non-falciparum species among clinical malaria patients.[23] There has been one recent report of *P. vivax* commonly occurring in Hue.[23] These inconsistent declines in prevalence suggest the need to characterize the epidemiology of asymptomatic and non-falciparum infections in this period to inform future control efforts.

We used survey, GPS, and biospecimen data collected in the 2014-15 Rwanda Demographic and Health Survey (RDHS), a large nationally population-representative study, to characterize asymptomatic and non-falciparum infections across the country’s adult population.

## Methods

### Ethics Statement

Dried blood spots (DBS) were provided by the Ministry of Health-Rwanda. This research was reviewed and approved by the Rwanda National Ethics Committee (reference 102/RNEC/2023). The University of North Carolina and Brown University IRBs deemed this non-human subjects research.

### Study design and population

The 2014-2015 Rwanda DHS studied 12,699 households from 492 GPS-located clusters from all 30 recognized districts. In 50% of households, a total of 16,930 dried blood spot (DBS) specimens were collected for HIV testing from men aged 15-59 years and women aged 15-49 years, and a subsample of children 0-14 years; in the other 50% of households, rapid malaria diagnostic testing was completed on children aged 6-59 months.[24] Previously, we used these data to estimate clusters that would represent high and low malaria transmission areas.[25] High prevalence clusters had a RDT or microscopy positivity rate of >15%. We included 1,887 samples from these 56 high prevalence clusters in 3 regions. In addition to samples from high prevalence areas, a random subset of 3,163 samples from 401 low prevalence clusters were selected. A total of 5,050 DBS samples from 457 out of 492 DHS clusters were analyzed for four species of malaria infection by real time PCR. (**S1 Figure**).

### Species-specific real time PCR

DNA from each sample was extracted from three 6 mm DBS punches using Chelex and screened for four species of malaria infection using real-time PCR assays.[26] These assays (**S1 Table**) targeted the 18s genes for *P. malariae, P. ovale*, and *P. vivax*, and the varATS repeat in *P. falciparum*.[20,27–29] To allow for quantification of *P. falciparum*, mock DBS were created using whole blood and cultured 3d7 parasites (MRA-102, BEI Resources, Manasas, VA) and extracted with the same assay used for samples. Controls for non-falciparum species used serial dilutions of plasmid DNA (MRA-180, MRA-179, MRA-178; BEI Resources, Manassas VA), with estimates for parasitemia based on an estimated six 18s rRNA gene copies per parasite.[30] All assays were run for 45 cycles to enable detection of lower density infections. The high cycle number approach has been previously evaluated for *P. ovale* and *P. vivax*, where assays were tested against 390 negative controls (human DNA) with no false positives.[31] We had no false positive results in our non-template controls for this study. In addition, we ran each assay to 45 cycles against plasmids of the other species to evaluate for cross-reactivity (**S2 Table**). All positive samples were confirmed by manually reviewing the amplification curves in the machine software. Standard curves had a minimum r-squared value of 0.95 across all runs. A positive result for each species was determined using a 45-cycle cutoff, unless otherwise stated. In addition, the assays retain a high specificity at high cycle number.[32]

### Spatial & ecological variables

Deidentified survey and geospatial data from the 2014-2015 Rwanda DHS were matched to PCR data using DBS sample barcodes. Clusters with individuals positive for any species of malaria infection were mapped using DHS geospatial coordinates. Individual level covariates assessed for association included sex, age group, wealth quintile, education level, livestock ownership, source of drinking water, bed net ownership, whether the household bed net has been treated with long-lasting insecticide (LLIN = long-lasting insecticide-treated net) and sleeping under a LLIN the night prior to the survey. Cluster level covariates included region, urban/rural status of place of residence, elevation, month of data collection, proportion of a given cluster living in a household with a bed net, proportion of the cluster that slept under an LLIN, land cover, average daily maximum temperature for the current month and precipitation for the prior month. Land cover estimates were taken from the Regional Center for Mapping Resources for Development and SERVIR-Eastern and Southern Africa, cluster-level temperature values were obtained from the 2014-2015 Rwanda DHS geospatial covariates, and precipitation values from the Climate Hazards Group InfraRed Precipitation with Station (CHIRPS) dataset. [33] Survey clusters were assigned GPS coordinate values within buffers as described previously in accordance with DHS specifications.[31,34]

### Statistical analysis

We estimated species-specific prevalence, non-falciparum prevalence, and overall *Plasmodium sp*. prevalence, applying HIV sampling weights, inverse propensity for selection weights, and weights to account for selection by cluster transmission intensity and the skewed selection of samples from low and high transmission clusters. [35,36] HIV sampling weights are provided in DHS data and account for potential selection bias resulting from differential DBS collection among survey participants. The DHS statistics guide recommends inclusion of individual level sampling weights if participants are sampled in a structured way (over-sampling from high transmission clusters). These three weights were multiplied for the final weights used for each participant included in the analysis. We estimated bivariate associations between each *Plasmodium* species and a variety of covariates available in the DHS and investigated in other contexts,[31,37] including household wealth, livestock ownership, or bed net use, using the same combination of weights. We report prevalence differences and 95% confidence intervals to assess precision. Multivariable analysis was not performed as the combination of weights applied should produce estimates representative of the full survey and country. We analyzed data using the *survey* (4.2.1), *srvyr* (v1.2.0), and *sf* (v1.0-8) packages using R 4.2.1 (R Foundation for Statistical Computing). Shapefiles of Rwanda district boundaries and water bodies were downloaded from the OCHA Regional Office for Southern and Eastern Africa database, and borders for neighboring countries were downloaded from ArcGIS Hub.[38] [39] [40]

## Results

### Study population characteristics

The study population (n = 5,050, weighted n = 5,099) was 51% (2,609/5,099) female, 32% (1,651/5,099) aged 15-24 years, 76% (3,538/5,099) lived in rural areas, and 75% (3,831/5,099) had a primary school education or no education (63% (3,191/5,099) reported primary education, 13% (640/5,099) reported preschool/none). Only 1.5% (77/5,099) of the study population was aged 0-14 years due to limited sampling of children in the original DHS, so the results of this survey are only representative of Rwandan adults. Overall, most (82% (4,196/5,099)) individuals reported a household bed net and 62% (3,173/5,099) reported sleeping under a long-lasting insecticide treated net the night before the survey. However, 76% (3,880/5,099) of the study population lived in a household that did not meet the World Health organization’s criteria of at least 1 net per 1.8 household members. For each of our covariates of interest, the study population was mostly comparable to and representative of the overall DHS population (**S3 Table**).

### Prevalence of P. falciparum and non-falciparum infection by real time PCR

A total of 1,478 *P. falciparum*, 283 *P. ovale*, 186 *P. malariae* and 7 *P. vivax* infections were identified. The overall weighted prevalence of any non-falciparum malaria infection was 8.0% (407/5,099, 95% CI: [6.6, 9.3]) compared to 16.3% (831/5,099 [14.5, 18.1]) for *P. falciparum* and 22.3% (1,137/5,099 [20.3, 24.3]) overall malaria prevalence with HIV sampling, inverse propensity of selection, and transmission intensity correction weights applied. Species specific weighted prevalences were 3.4% (172/5,099) [2.6, 4.1] and 4.7% (241/5,099) [3.5, 5.9] for *P. malariae* and *P. ovale spp*. Unweighted prevalence for *P. vivax* was 0.14%, (7/5,050) with a manually calculated 95% CI [0.03, 0.25]. Study population characteristics and the distribution of PCR positives by species are shown in **Table 1**. Unweighted prevalence estimates were calculated for each species based on false-positive rates of high cycle number PCR (**S4 Table**). Using a more restrictive cut off of 40 cycles (requiring a higher parasitemia to be positive at approximately 1 parasite per microliter of template DNA), resulted in weighted overall prevalences of 4.3% (218/5,099) [3.5, 5.1] for any non-falciparum infection, compared to 13.2% (674/5,099) [11.6, 14.8] for *P. falciparum* and 16.4% (836/5,099) [14.7, 18.1] for overall malaria. Species specific weighted prevalences at this cut off were 2.7% (139/5,099) [2.0, 3.4] and 1.6% (80/5,099) [1.1, 2.0], and for *P. malariae*, and *P. ovale spp*. Unweighted prevalence for *P. vivax* was 0.06% (3/5,050 [0.00, 0.13]. The largest difference in estimated prevalence was for *P. ovale spp*. This is not surprising given the distribution of estimated parasitemia values (**Figure 1**), showing a lower median parasitemia in the non-falciparum species compared to *P. falciparum*. District level weighted prevalences and their differences by PCR cut off are shown in **S5 Table** and **S6 Table**. *P. falciparum, P. ovale* and *P. malariae* infections were distributed across the country, while *P. vivax* infections were more localized (**S2 Figure**). District level overall malaria prevalence is shown in **Figure 2**, while district level prevalences for each species are illustrated in **Figure 3**. Among *P. ovale spp. P. malariae*, and *P. vivax* infections, 48.4%, (137/283) [42.5, 54.4]), 46.8% (87/186) [39.5,54.2]), and 57% (4/7) [20.2, 88.2], (unweighted counts) were infected with at least one other species of malaria (**S7 Table**).

**Table 1.**
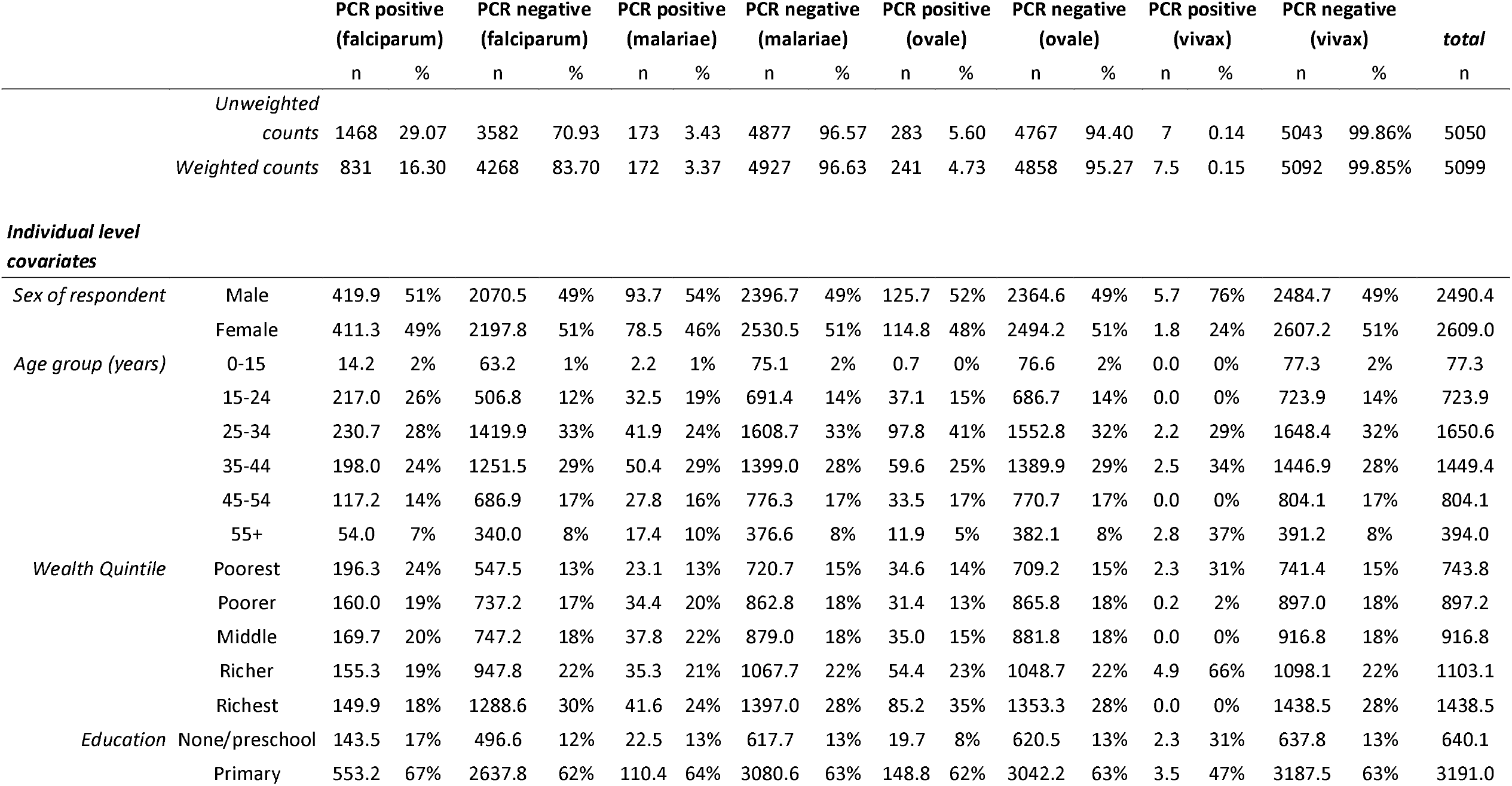
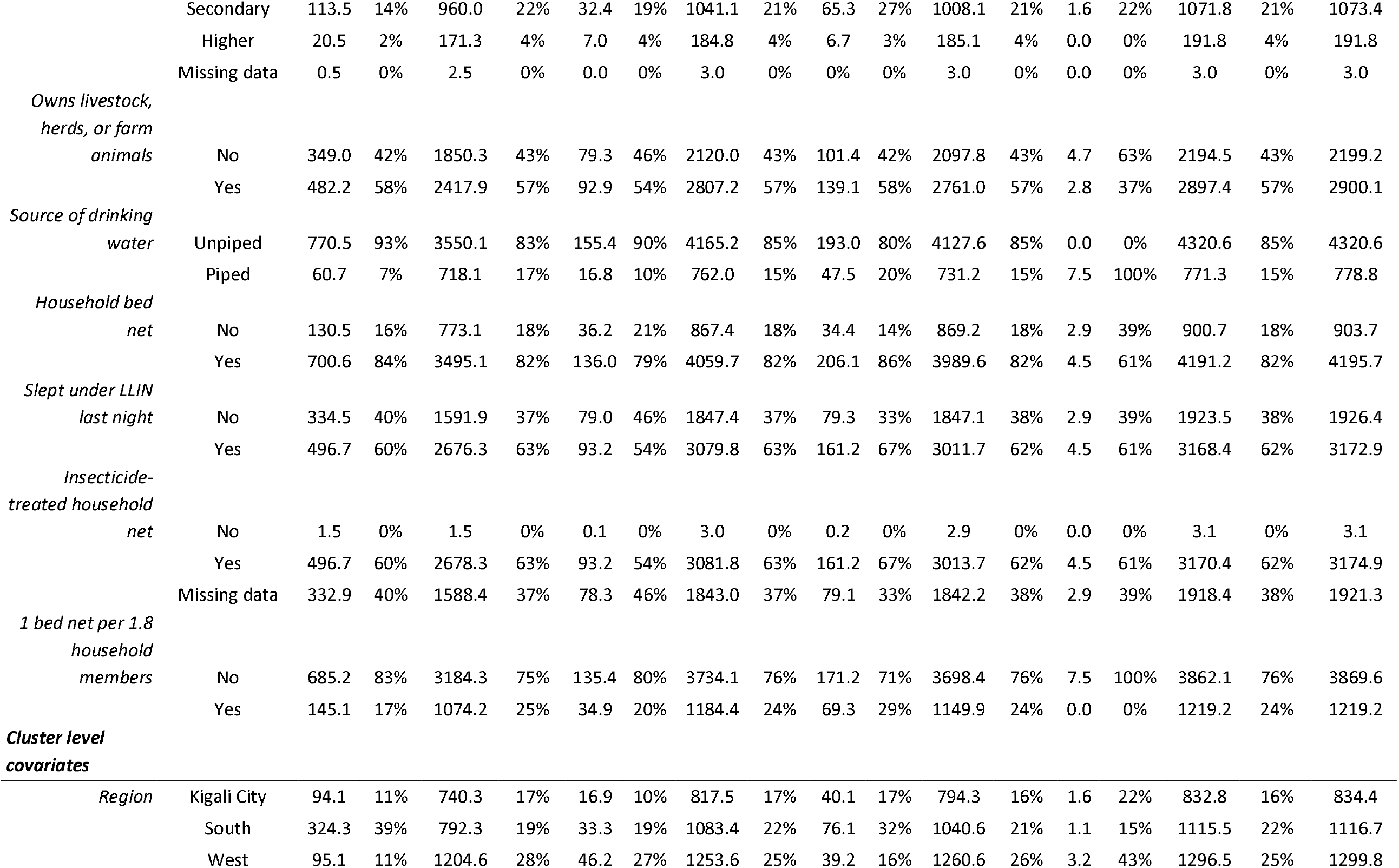
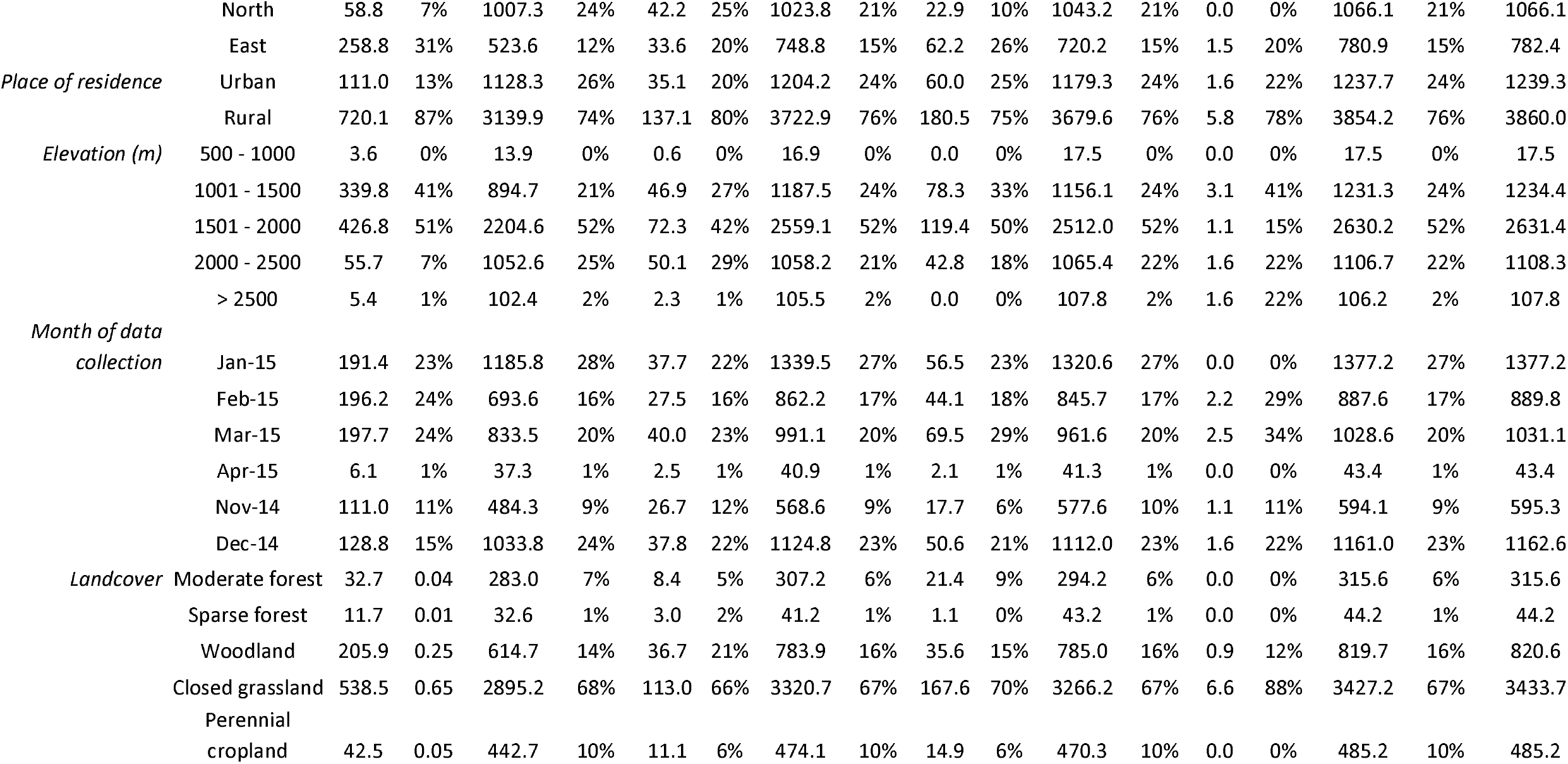
Study Population Characteristics and Distribution of PCR Positives by Species. Weighted counts and percentages for all individuals positive for each species of malaria. Weights applied are DHS sampling weights, propensity score weights, and weights to correct for over-selection of high transmission intensity clusters (transmission intensity weights). Characteristics of the included study population are also shown.

**Figure 1.**
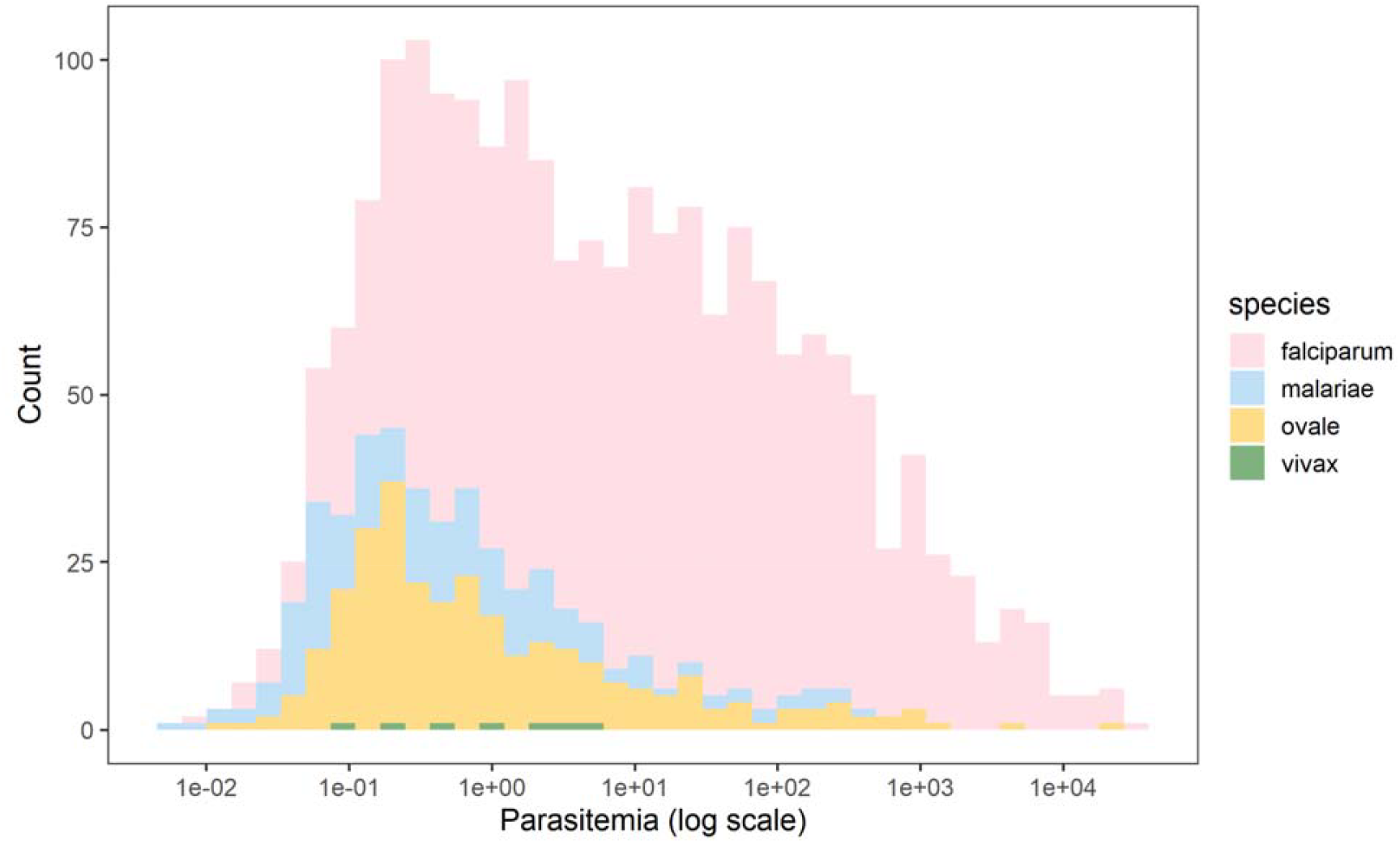
Calculated Parasitemia Estimates for Falciparum and Non-falciparum Infections. Overall, falciparum had a higher parasite density with a median of 10.90 (IQR 0.96-101.41). The median parasitemia level for *P. malariae, P. ovale spp*., *and P. vivax malaria were 0.35 (IQR: 0.07-1.83), 0.48 (IQR: 0.16-3.25) and 0.94 (IQR: 0.20-3.44), respectively*.

**Figure 2.**
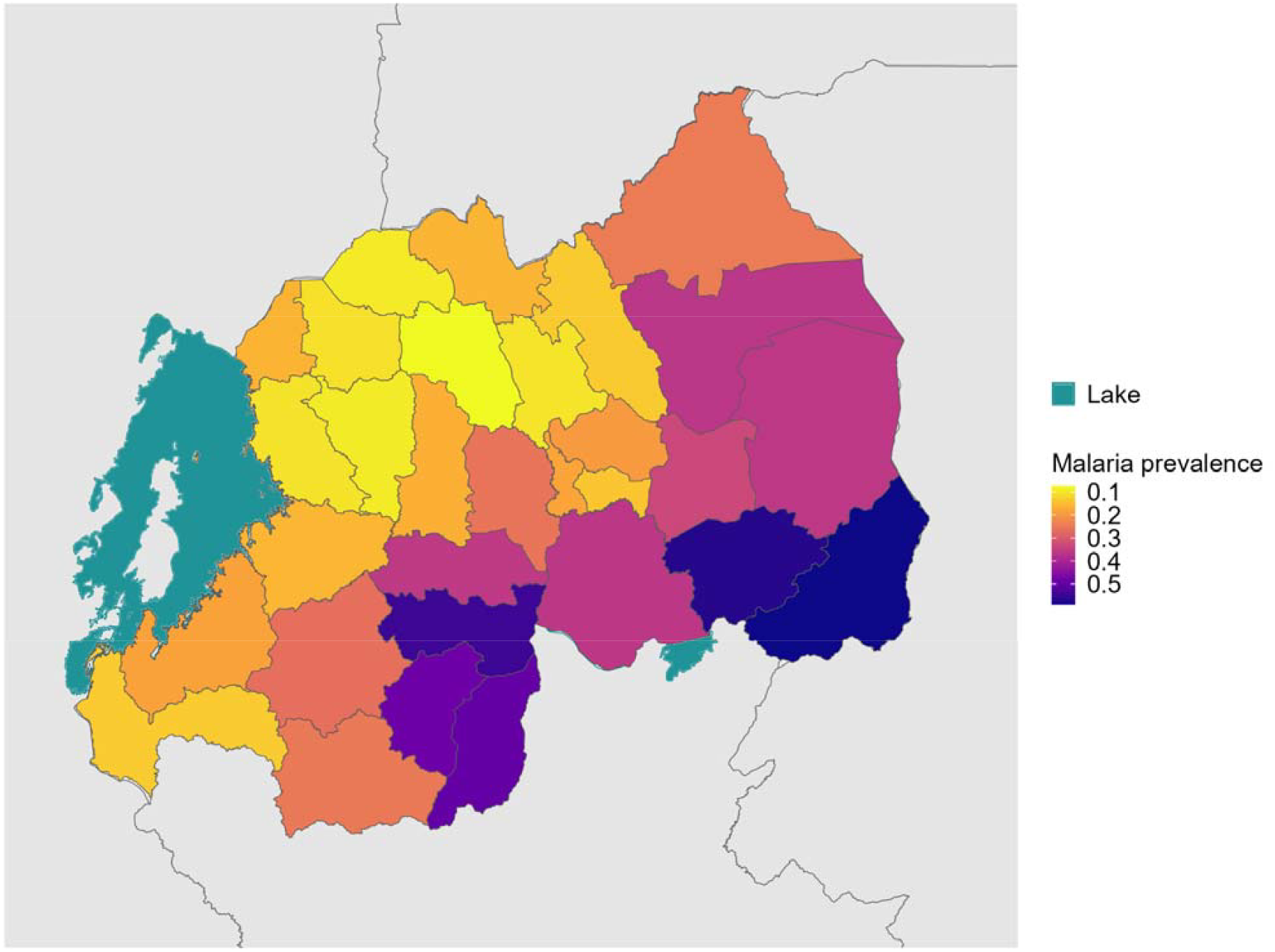
District Level Weighted Overall Malaria Prevalence. Weighted prevalence of any malaria infection, using HIV sampling, inverse propensity for selection and transmission intensity weights (described above). Shapefiles for country borders were downloaded from ArcGIS Hub, district boundaries and the lake shapefile were downloaded from the OCHA Regional Office for Southern and Eastern Africa database.

**Figure 3.**
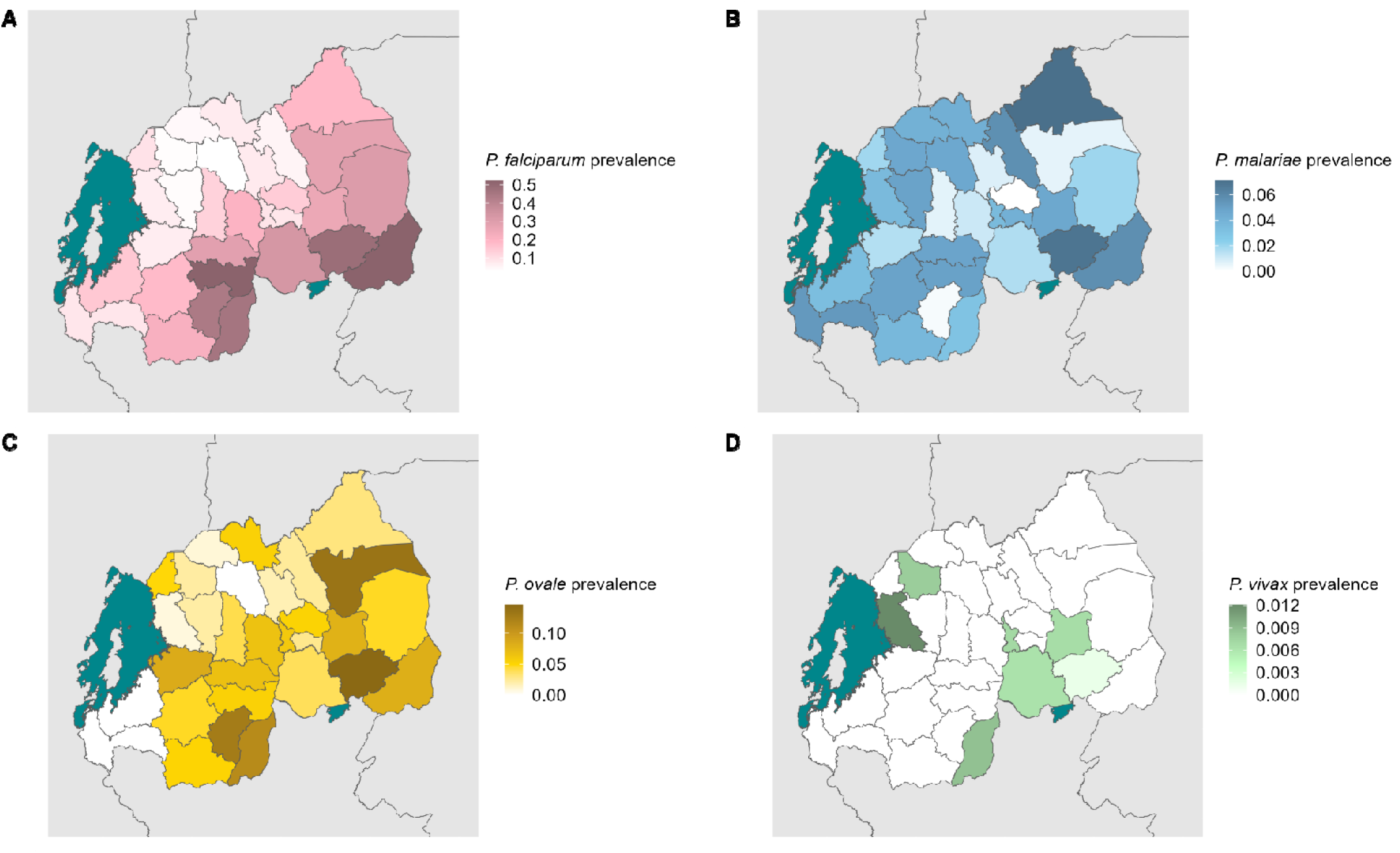
District Level Weighted Prevalence Estimates for Malaria Species. The weighted prevalence estimate for each species is shown. Panel A, B, C and D represent *P. falciparum, P. malariae, P. ovale spp. and P. vivax* malaria, respectively. Shapefiles for country borders were downloaded from ArcGIS Hub, district boundaries and the lake shapefile were downloaded from the OCHA Regional Office for Southern and Eastern Africa database.

### Bivariate associations for infection

Bivariate regression models using weighted (as previously described) survey data found multiple associations for infection with *P. falciparum* malaria, but few for non-falciparum malaria (**Figure 4**). Covariates with more than two categories were dichotomized to provide interpretable comparisons across the study population. For example, Anopheles mosquito vectors generally do not thrive at elevations over 1,500 meters, although this is not a fixed limit. [41] Land cover classifications were dichotomized to compare less vegetation/more human activity to more vegetation/less human activity. No adjusted analyses were conducted as we are only exploring the strength of individual associations based on prevalence measures, and generalizability of participants analyzed to the country or survey as a whole is provided through weighting. Similar to previous work, a higher prevalence of *P. falciparum* malaria in our dataset was significantly (at a 0.05 confidence level for all associations) associated with multiple study covariates related to socioeconomic status (lower wealth quintile, lower education status, and unpiped drinking water). Secondary or higher education, residence in a household with at least 1 bed net per 1.8 household members, piped drinking water, continuous increase (and clusters at or above the study sample average) in prior month’s average rainfall, and higher altitude (>1,500 meters) were all associated with significantly lower prevalence of *P. falciparum* malaria. Rural clusters compared to urban, clusters designated as perennial cropland or grassland (compared to woodland or sparse forest), lower wealth index (first and second quintiles compared to the upper three), younger age (15-24 years compared to over 24 years) and residence in clusters with monthly average temperatures at or above the average for all clusters in analysis were associated with higher *P. falciparum* prevalence. Fewer associations were found for non-falciparum malaria. No significant associations were found for P. malariae infection at an alpha level of 0.05. *P. ovale* infection was significantly associated with residence in clusters at or above the average monthly temperature, while lower prevalence of infection was associated with younger age in both the binary categorization described above and continuous decrease in age. Associations for *P. vivax* infection were not attempted due to the limited number of infections in the survey (n=7).

**Figure 4.**
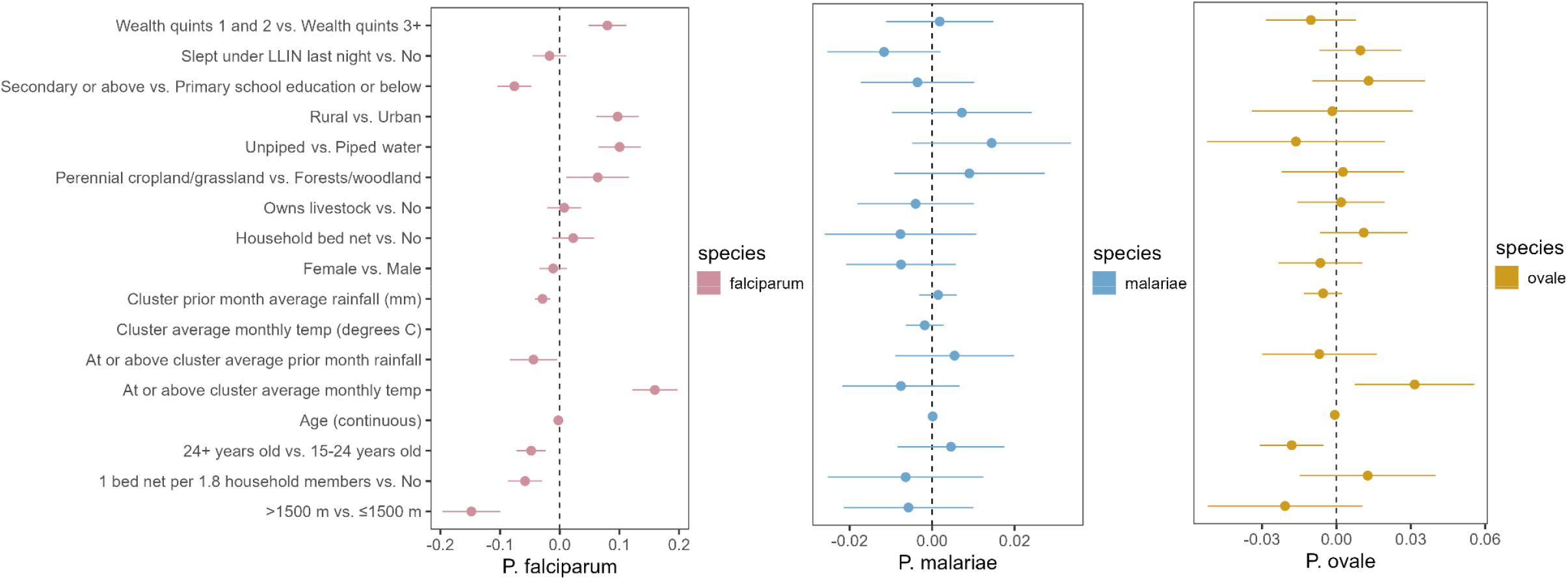
Bivariate associations and between demographic and environmental risk factors and *Plasmodium* spp. prevalence using weighted survey data. Models incorporate 2014-15 Rwanda Demographic and Health Survey weights, inverse probability of selection weights, and cluster transmission intensity weights (described in **Table 1**). Point estimates of prevalence difference are surrounded by confidence intervals. The reference is the second variable listed. Panel A, B and C represent *P. falciparum, P. malariae, and P. ovale sp*., respectively. Note that each panel has a different scale.

## Discussion

In Rwanda, a setting with robust malaria control efforts but inconsistently declining prevalence, we conducted the largest assessment of asymptomatic and non-falciparum malaria to-date, primarily among adults. We found an asymptomatic malaria weighted prevalence of 22.3%, using PCR, substantially higher than estimates produced using RDT and microscopy. Additionally, we observed 8.0% of the population was infected with a non-falciparum species. While we observed *P. falciparum* prevalence to be 16.3% in adults, malaria detection by microscopy in the DHS reported a 2% malaria prevalence among children age 6-59 months and 0.6% among women age 15-49.[42] This high prevalence in adults found using molecular detection is consistent with a study of school-aged individuals in the Huye District in the same year, finding a 22% prevalence of any malaria (19% *P. falciparum* prevalence) using a combined microscopy and PCR approach.[43]

We used an ultra-sensitive assay for *P. falciparum* to detect primarily asymptomatic, lower density infections, improving the characterization of *P. falciparum* prevalence. While a higher PCR cycle cut-off may cause concern about false-positive detection, our assays have been run extensively at 45 cycles with little to no evidence of false positives[31], and are consistent with estimates using a lower cut off (40 cycles). The largest change in prevalence occurred with *P. ovale spp*., where estimated prevalence dropped from 4.7% to 1.6% at different cycle cut offs, reflecting the high number of low density infections detected. The relative decrease in prevalence for each species based on cycle cutoff was not always consistent, with some regions having no decline in *P. falciparum* prevalence with large declines in *P. ovale* malaria (e.g. Karongi near Lake Kivu) or the opposite with no decline in non-falciparum but lower *P. falciparum* prevalence estimates (e.g. Rutsiro and Nyamasheke). With asymptomatic *P. falciparum* malaria increasing risk of symptomatic disease at one month and contributing to significant morbidity, the high observed prevalence of asymptomatic, low-parasitemia infections offers an important direction for malaria control.[10,11] Estimating prevalence of sub-patent malaria should be prioritized as a complement to RDT or microscopy detected prevalence to accurately characterize transmissible reservoirs in endemic regions.

Non-falciparum malaria was detected in 8.0% of individuals nationally, a prevalence not previously appreciated in the country. *P. ovale spp*. and *P. malariae* were both common in Rwanda (4.7% and 3.3% prevalence, respectively) and distributed in regions of both high and low transmission. A recent household survey estimated that *P. falciparum* is responsible for 97% of malaria infections in Rwanda, with *P. malariae* and *P. ovale spp*. each responsible for 1%–2% of total infections (Uwimana A. {Unpublished]).[44] *P. vivax* is present but remains relatively uncommon (0.14% unweighted prevalence) and sporadic, but consistent with reporting of occasional clinical cases. [44] More recently, a cluster of *P. vivax* was reported in the Huye District.[23] Given distinct treatment approaches by species, the higher than expected prevalence of *P. ovale spp*. and occasional *P. vivax* infection underscores the need for molecular monitoring to guide control efforts towards elimination. Regular surveillance for non-falciparum malaria is needed to inform regional treatment guidelines, particularly if relapsing malaria species continue to expand their prevalence.

Not surprisingly, mixed species infections were common and widely distributed, but occurred more commonly in clusters in the south and east where malaria transmission was the highest (**S2 Figure**). Among *P. ovale spp. P. malariae*, and *P. vivax* infections, 44%, 45% and 57% (unweighted) were infected with at least one other species of malaria (**S7 Table**). Bivariate associations for mixed infections largely reflected those observed for *P. falciparum*, albeit with lower magnitude and wider confidence intervals attributable to the smaller total counts. Mixed infections are often underappreciated by clinicians and laboratorians and may lead to severe disease complications. A recent meta-analysis suggested that patients with mixed infections have a higher proportion of pulmonary complications and multiple organ failure than patients with *P. falciparum* infection alone.[45] The impact of mixed species infections on clinical malaria outcomes in Rwanda is unknown and requires additional evaluation in symptomatic infections, which were not included in this study.

The associations we observed between *P. falciparum* infection and lower household wealth, no bed net use and lower elevation are consistent with previous studies using these data, with malaria positivity defined by RDT or microscopy.[46,47] We observed malaria prevalence was highest in the South and East as expected, but asymptomatic infection remained common in other areas. Like similar studies, associations of covariates for non-falciparum malaria were few and traditional risk factors for *P. falciparum* were less strongly associated with non-falciparum malaria.[20,28,31,37] This raises concern for how the control program can target non-falciparum infections without better diagnostics in the community. The reasons for the relative lack of risk factors, especially for *P. ovale spp*., remains unclear. Relapsing malaria, caused by *P. ovale* and *P. vivax*, may not be associated with typical covariates due to the inability to discern between incident or relapse infections in the study. Additionally, non-falciparum infections are under-detected in this sample compared to falciparum due to the ultra-sensitive assay used for falciparum detection, which may bias estimates and increase uncertainty for associations with non-falciparum infections.

While this data reflects the epidemiology from nearly a decade ago, it remains important for malaria control. The baseline set here is useful for understanding the interventions that have been used in Rwanda for malaria control if future similar surveys, such as the 2019/20 DHS, Malaria Indicator Surveys, or other broad sampling efforts, are genotyped similarly. The longitudinal data across surveys can be used to understand how malaria interventions impact both asymptomatic malaria and non-falciparum malaria, both of which are not captured by standard DHS data but still important for malaria elimination.

While this national molecular evaluation offers important perspectives, we are limited by a few key points. As noted above, the cross-sectional nature of a single DHS limits inference regarding transmission and time trends, but use of subsequent (2019-20 DHS) and future DHSs, would allow for ongoing surveillance of these parasites, as has been done in the DRC.[48] We also could not determine if *P. ovale* spp. and *P. vivax* infections were newly acquired or the result of relapse from hypnozoites. Detection of these parasites is still important for malaria control programming. Additionally, the relative lack of data for children under 15 makes this survey not representative of the full asymptomatic reservoir and non-falciparum prevalence. We are limited by the sampling framework of the DHS. School aged children are a particularly vulnerable group and often have the highest rate of malaria infection in Africa, including for *P. vivax*.[49] Without this group, a true population prevalence of asymptomatic infection is difficult to determine. However, we likely underestimate overall population prevalence given the 2017 MIS had an overall prevalence of 14% for children aged 6 months to 14 years of age using a rapid diagnostic test which typically detects 41% of infections compared to PCR.[9,50] In addition, males were overrepresented in the sample. We used an assay that targets 18S ribosomal RNA genes for non-falciparum species, thus could underreport infections that could have been detected with assays that target genes with higher copy numbers in the parasite such as the assay used for falciparum in this study. [51–53] Despite these limitations, the use of existing samples and individual level data from a DHS is a highly informative method to gain insights into national malaria prevalence.

This study represents the first nationwide molecular investigation of asymptomatic *Plasmodium falciparum* and non-falciparum malaria in Rwanda and highlights a significant parasite reservoir and the essential role of molecular testing to appropriately detect them. First, we found that asymptomatic *P. falciparum* infections in adults were significantly more prevalent than previously estimated by rapid diagnostic tests (RDTs) and microscopy in children.[42] These infections are not benign: they carry clinical consequences and serve as a hidden reservoir for continued transmission, underscoring the importance of identifying and addressing asymptomatic carriers in control strategies.[10,12] Second, molecular methods were necessary to detect *P. ovale spp*., *P. malariae*, and *P. vivax*, which are better characterized by molecular methods as they can be frequently missed by standard diagnostics. In this study, most non-falciparum infections were low-density and often co-occurred with *P. falciparum*, particularly *P. ovale spp*. and *P. vivax*, which would contribute to under-recognition of mixed infections. The observed prevalence of *P. ovale spp*. and *P. malariae* was higher than expected and lacked consistent risk factors. In order to assess if these species may be expanding into niches left by successful *P. falciparum* control, continued monitoring with molecular methods is needed. Third, comprehensive malaria elimination efforts must address all human malaria species—not just *P. falciparum*—including those capable of relapse and silent transmission. Therefore, ongoing molecular surveillance, ideally led by the national malaria control program, is critical to track changing epidemiology, guide targeted interventions, and support Rwanda’s path toward malaria elimination.

## Author Contributions

CK, CM, JAB, JBM and JJJ conceived of the study. CG, CM, RK, SW, HMT, DG, KT, and KB were involved in data generation and analysis. All authors were involved in writing the manuscript.

## Data Availability

Clinical and spatial data are available through the DHS MEASURE website (https://dhsprogram.com/data/available-datasets.cfm). PCR data has been provided to DHS and is available upon request with approval of use of DHS data from DHS MEASURE. Code used for analysis is available at: https://github.com/claudiagaither.

## Acknowledgements

The following reagent was obtained through BEI Resources, NIAID, NIH: *Plasmodium falciparum*, Strain 3D7, MRA-102, contributed by Daniel J. Carucci. The following reagent was obtained through BEI Resources, NIAID, NIH: Diagnostic Plasmid Containing the Small Subunit Ribosomal RNA Gene (18S) from *Plasmodium vivax*, MRA-178, contributed by Peter A. Zimmerman. The following reagent was obtained through BEI Resources, NIAID, NIH: Diagnostic Plasmid Containing the Small Subunit Ribosomal RNA Gene (18S) from *Plasmodium malariae*, MRA-179, contributed by Peter A. Zimmerman. The following reagent was obtained through BEI Resources, NIAID, NIH: Diagnostic Plasmid Containing the Small Subunit Ribosomal RNA Gene (18S) from *Plasmodium ovale*, MRA-180, contributed by Peter A. Zimmerman.

