## Supplementary material for "Prevalence of Asymptomatic non-Falciparum and Falciparum Malaria in the 2014-15 Rwanda Demographic Health Survey": Figure S1

**S1 Figure. Distribution of 2014-2015 Rwanda Demographic Health Survey Clusters.** Clusters are color coded based on malaria transmission intensity, based upon DHS malaria testing results (high transmission represents >15% positive by RDT or microscopy. Shape fill is based on the cluster's inclusion in this analysis. Shapefiles for country borders were downloaded from ArcGIS Hub, the lake shapefile from the OCHA Regional Office for Southern and Eastern Africa database.

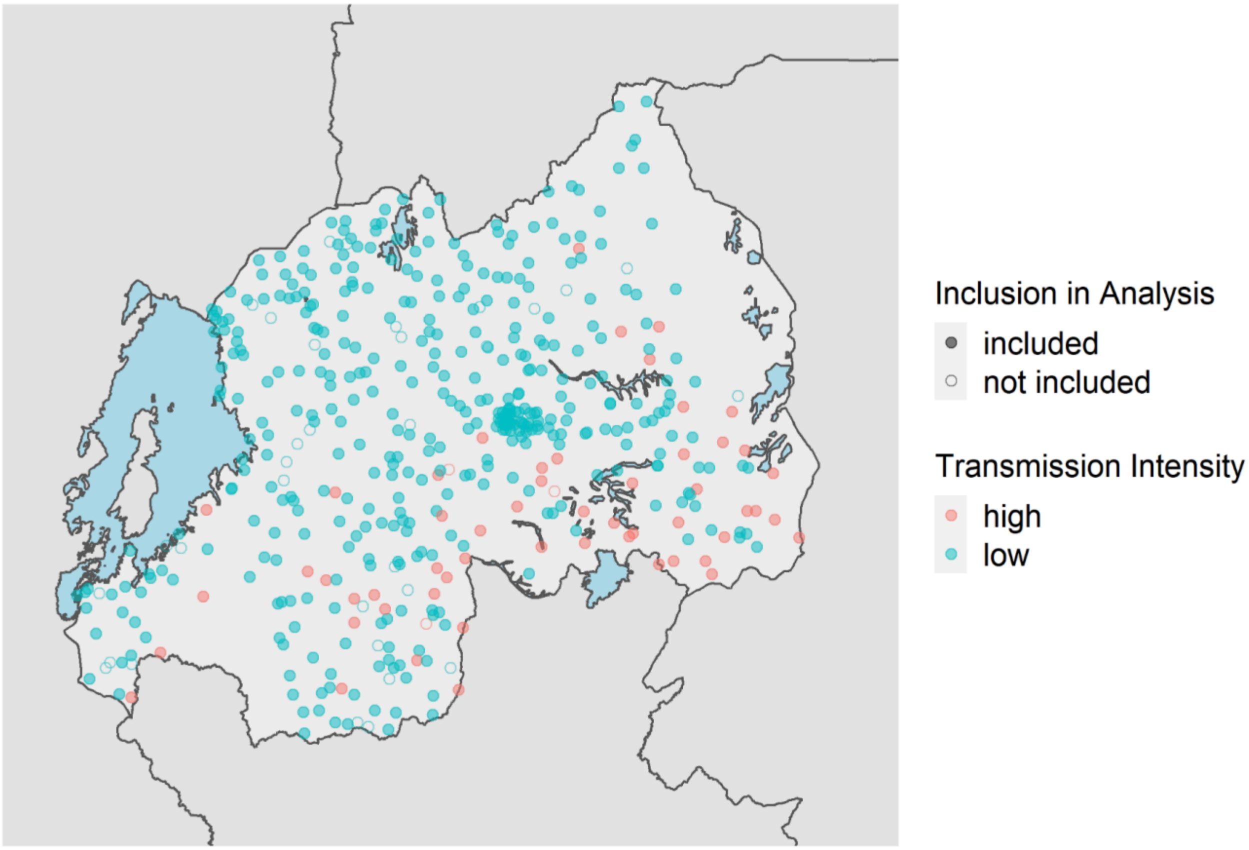
