## Supplementary material for "Prevalence of Asymptomatic non-Falciparum and Falciparum Malaria in the 2014-15 Rwanda Demographic Health Survey": Figure S2

**S2 Figure. Distribution of clusters where malaria species were Identified.** Mixed infections and mono infections are differentiated by cluster shape and color. Shapefiles for country borders were downloaded from ArcGIS Hub, the lake shapefile from the OCHA Regional Office for Southern and Eastern Africa database.

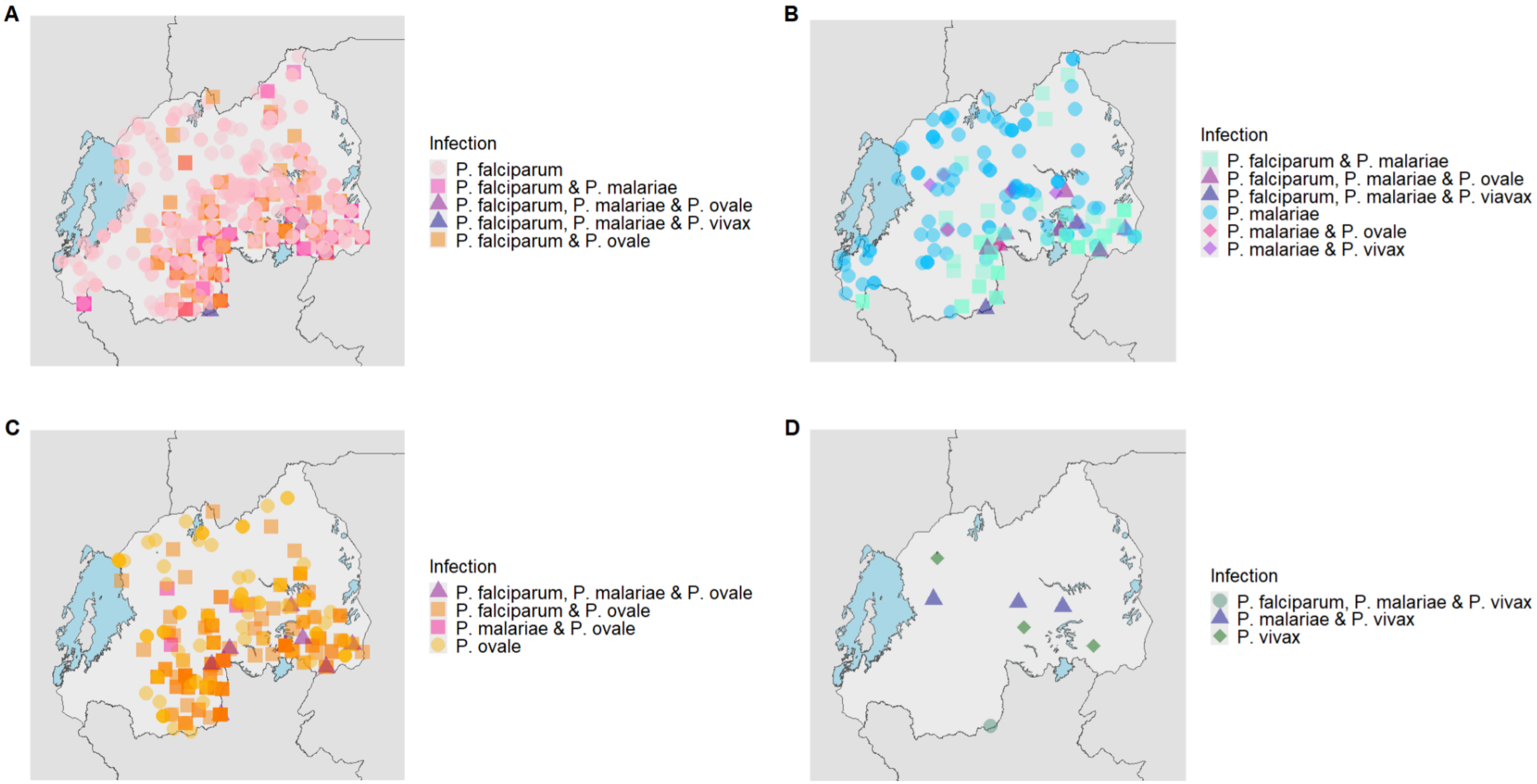
