## Supplementary material for "Prevalence of Asymptomatic non-Falciparum and Falciparum Malaria in the 2014-15 Rwanda Demographic Health Survey": Table S1

**S1 Table. PCR Primers, Probes and Reaction Conditions.** Malaria speciation qPCR assay details.

### *Plasmodium falciparum (varATS)*

*Adapted from:* Hoffman, N, et. al. PLOS Medicine. 2015.

*Forward Primer* 5'- CCCATACACAACCAAYTGGA - 3'
*Reverse Primer* 5' - TTCGCACATATCTCTATGTCTATCT - 3'
*Probe* 5' - 6-FAM-TRTTCCATAAATGGT-NFQ-MGB - 3'

| *Cycling conditions* | | | 45 cycles | |
|---|---|---|---|---|
| *temp (degrees Celsius)* | 50 | 95 | 95 | 55 |
| *time* | 2 min | 10 min | 15 s | 1 min |

| *Roche FastStart Universal Probe Master* | |
|---|---|
| *Fwd primer* | 800 nM |
| *Rev primer* | 800 nM |
| *Probe* | 400 nM |
| Template DNA | 2.5 µl |
| Total volume | 12.5 µl |

### *Plasmodium malariae (18s)*

Rougemont M, et. al. Journal of Clinical Microbiology. 2004.

5' - AGTTAAGGGAGTGAAGACGATCAGA - 3'
5' - CAACCCAAAGACTTTGATTTCTCATAA - 3'
5' - 6-FAM-ATGAGTGTTTCTTTTAGATAGC-NFQ-MGB - 3'

| | | | 45 cycles | |
|---|---|---|---|---|
| | 50 | 95 | 95 | 60 |
| | 2 min | 10 min | 15 s | 1 min |

| *Roche FastStart Universal Probe Master* | |
|---|---|
| *Fwd primer* | 300 nM |
| *Rev primer* | 300 nM |
| *Probe* | 400 nM |
| Template DNA | 2 µl |
| Total volume | 12.5 µl |

### *Plasmodium ovale (18s)*

*Adapted from:* Mitchell C, et. al. Journal of Infectious Diseases. 2021

*Forward Primer* 5’ - CCRACTAGGTTTTGGATGAAAVRTTTTT- 3’
*Reverse Primer* 5’ - AACCCAAAGACTTTGATTTCTCATAA - 3’
*Probe* 5’ - VIC/CRAAAGGAATTYTCTTATT - 3’

| *Cycling conditions* | | | 45 cycles | |
|---|---|---|---|---|
| *temp (degrees Celsius)* | 50 | 95 | 95 | 52 |
| *time* | 2 min | 10 min | 15 s | 1 min |

| *Roche FastStart Universal Probe Master (Rox)* | |
|---|---|
| *Fwd primer* | 400 nM |
| *Rev primer* | 400 nM |
| *Probe* | 200 nM |
| Template DNA | 2 µl |
| Total volume | 12 µl |

### *Plasmodium vivax (18s)*

Brazeau N, et. al. Nature Communications. 2021

5’ - ACGCTTCTAGCTTAATCCACATAACT - 3’
5’ - ATTTACTCAAAGTAACAAGGACTTCCAAGC - 3’
5' - /56-FAM/TTCGTATCG/ZEN/ACTTTGTGCGCATTTTGC/3IABkFQ/ - 3'

| | | | 45 cycles | |
|---|---|---|---|---|
| | 50 | 95 | 95 | 60 |
| | 2 min | 10 min | 15 s | 1 min |

| *Roche FastStart Universal Probe Master (Rox)* | |
|---|---|
| *Fwd primer* | 400 nM |
| *Rev primer* | 400 nM |
| *Probe* | 200 nM |
| Template DNA | 5 µl |
| Total volume | 18 µl |
