## Supplementary material for "Prevalence of Asymptomatic non-Falciparum and Falciparum Malaria in the 2014-15 Rwanda Demographic Health Survey": Table S2

**S2 Table. Cross reactivity of non-falciparum real time PCRs at 45 cycles.** Each assay was run to 45 cycles against plasmids diluted to representative parasitemias (based on 6 copies per genomic equivalent).

| *Assay* | *Template* | *Genomic equivalent (GE)* | *# positive* | *CT of positive* |
|---|---|---|---|---|
| *P. vivax* | *P. falciparum* | 212 GE/µL | 0/20 | NA |
| | *P. ovale* | 110 GE/µL | 0/20 | NA |
| | *P. malariae* | 100 GE/µL | 1/20 | 42.1 |
| *P. ovale* | *P. falciparum* | 212 GE/µL | 1/20 | 43.7 |
| | *P. vivax* | 76.5 GE/µL | 0/20 | NA |
| | *P. malariae* | 100 GE/µL | 1/20 | 42.5 |
| *P. malariae* | *P. falciparum* | 212 GE/µL | 0/20 | NA |
| | *P. vivax* | 76.5 GE/µL | 0/20 | NA |
| | *P. ovale* | 110 GE/µL | 0/20 | NA |
