## Supplementary material for "Prevalence of Asymptomatic non-Falciparum and Falciparum Malaria in the 2014-15 Rwanda Demographic Health Survey": Table S3

**S3 Table. Comparison of Population Used for Molecular Screening to Overall DHS Population.** Weighted counts in each category of analysis variables, for both the study population and all participants with DBS collected for HIV testing in the 2014-15 DHS.

| *Variable* | | *This Study* | *%* | *Total DHS HIV DBS* | *%* |
|---|---|---|---|---|---|
| *Sex of respondent* | Male | 2490 | 48.84% | 8314 | 49.02% |
| | Female | 2609 | 51.16% | 8645 | 50.98% |
| *Age group (years)* | 0-14 | 77 | 1.52% | 230 | 1.36% |
| | 15-24 | 724 | 14.20% | 4363 | 25.73% |
| | 25-34 | 1651 | 32.37% | 4758 | 28.05% |
| | 35-44 | 1449 | 28.42% | 4138 | 24.40% |
| | 45-54 | 804 | 15.77% | 2340 | 13.80% |
| | 55+ | 394 | 7.73% | 1129 | 6.66% |
| *Wealth Quintile* | Poorest | 744 | 14.59% | 2891 | 17.05% |
| | Poorer | 897 | 17.59% | 3245 | 19.14% |
| | Middle | 917 | 17.98% | 3418 | 20.16% |
| | Richer | 1103 | 21.63% | 3562 | 21.01% |
| | Richest | 1439 | 28.21% | 3841 | 22.65% |
| *Education* | None/preschool | 640 | 12.55% | 3262 | 19.23% |
| | Primary | 3191 | 62.58% | 10420 | 61.45% |
| | Secondary | 1073 | 21.05% | 2803 | 16.53% |
| | Higher | 192 | 3.76% | 456 | 2.69% |
| | Missing data | 3 | 0.06% | 17 | 0.10% |
| *Owns livestock, herds, or farm animals* | No | 2199 | 43.13% | 7279 | 42.92% |
| | Yes | 2900 | 56.87% | 9679 | 57.08% |
| *Source of drinking water* | Unpiped | 4321 | 84.73% | 15080 | 88.92% |
| | Piped | 779 | 15.27% | 1879 | 11.08% |
| *Household bed net* | No | 904 | 17.72% | 2802 | 16.52% |
| | Yes | 4196 | 82.28% | 14157 | 83.48% |
| *Slept under LLIN last night* | No | 1926 | 37.78% | 6367 | 37.54% |
| | Yes | 3173 | 62.22% | 10592 | 62.46% |
| *Insecticide-treated household net* | No | 3 | 0.06% | 20 | 0.11% |
| | Yes | 3175 | 62.26% | 10603 | 62.52% |
| | Missing data | 1921 | 37.68% | 6336 | 37.36% |
| *1 bed net per 1.8 household members* | No | 3870 | 76.04% | 13426 | 79.34% |
| | Yes | 1219 | 23.96% | 3497 | 20.66% |
| ***Cluster level covariates*** | | | | | |
| *Region* | Kigali City | 834 | 16.36% | 2156 | 12.71% |

| | | | | | |
|---|---|---|---|---|---|
| | South | 1117 | 21.90% | 4075 | 24.03% |
| | West | 1300 | 25.49% | 3768 | 22.22% |
| | North | 1066 | 20.91% | 2723 | 16.06% |
| | East | 782 | 15.34% | 4237 | 24.98% |
| *Place of residence* | Urban | 1239 | 24.30% | 3163 | 18.65% |
| | Rural | 3860 | 75.70% | 13796 | 81.35% |
| *Elevation (m)* | 500-1000 | 17 | 0.34% | 62 | 0.36% |
| | 1001-1500 | 1234 | 24.21% | 5036 | 29.70% |
| | 1501-2000 | 2631 | 51.60% | 8914 | 52.56% |
| | 2001-2500 | 1108 | 21.73% | 2798 | 16.50% |
| | 2500 < | 108 | 2.11% | 149 | 0.88% |
| *Month of data collection* | 15-Jan | 1377 | 27.01% | 4004 | 23.61% |
| | 15-Feb | 890 | 17.45% | 3019 | 17.80% |
| | 15-Mar | 1031 | 20.22% | 3628 | 21.40% |
| | 15-Apr | 43 | 0.85% | 107 | 0.63% |
| | 14-Nov | 595 | 11.67% | 2426 | 14.30% |
| | 14-Dec | 1163 | 22.80% | 3775 | 22.26% |
| *Land cover* | Moderate forest | 316 | 6.19% | 960 | 5.66% |
| | Sparse forest | 44 | 0.87% | 263 | 1.55% |
| | Woodland | 821 | 16.09% | 3260 | 19.23% |
| | Closed grassland | 3434 | 67.34% | 11253 | 66.36% |
| | Perennial cropland | 485 | 9.51% | 1222 | 7.21% |
