## Supplementary material for "Prevalence of Asymptomatic non-Falciparum and Falciparum Malaria in the 2014-15 Rwanda Demographic Health Survey": Table S4

**S4 Table. Impact of potential false positivity on unadjusted prevalence.** 3/160 replicates were false positive across assays: 1.9% [95% CI: 0.3-5.4] Binomial exact method. All adjustments rounded up to the higher integer.

| *Number tested* | | *# CT ≤ 40* | *# CT >40* | *Unweighted prevalence* | *0.3% false positivity among CT >40* | *Adjusted unweighted prevalence* | *5.4% false positivity among CT>40* | *Adjusted unweighted prevalence* |
|---|---|---|---|---|---|---|---|---|
| 5,050 | *P. falciparum* | 1297 | 181 | 29.3% | 180 | 29.3% | 171 | 29.1% |
| 5,050 | *P. malariae* | 159 | 27 | 3.7% | 27 | 3.7% | 26 | 3.7% |
| 5,050 | *P. ovale* | 124 | 159 | 5.6% | 159 | 5.6% | 150 | 5.4% |
