## Supplementary material for "Prevalence of Asymptomatic non-Falciparum and Falciparum Malaria in the 2014-15 Rwanda Demographic Health Survey": Table S5

**S5 Table. Differences in District Level Prevalence by PCR Cutoff.** Change in district level prevalence using only PCR positive infections with CT values 40 or less, compared to 45 cycles.

| *District* | *malaria_prevΔ* | *pf_prevΔ* | *pm_prevΔ* | *po_prevΔ* | *pv_prevΔ* |
|---|---|---|---|---|---|
| Nyarugenge | 0.114 | 0.037 | 0.025 | 0.052 | 0.000 |
| Gatsibo | 0.105 | 0.040 | 0.000 | 0.104 | 0.000 |
| Ngoma | 0.099 | 0.058 | 0.002 | 0.087 | 0.001 |
| Burera | 0.091 | 0.043 | 0.020 | 0.028 | 0.000 |
| Huye | 0.090 | 0.073 | 0.002 | 0.037 | 0.000 |
| Gasabo | 0.084 | 0.032 | 0.000 | 0.052 | 0.000 |
| Rwamagana | 0.084 | 0.052 | 0.014 | 0.063 | 0.000 |
| Kirehe | 0.082 | 0.052 | 0.002 | 0.043 | 0.000 |
| Kayonza | 0.074 | 0.060 | 0.000 | 0.021 | 0.000 |
| Ruhango | 0.072 | 0.026 | 0.030 | 0.046 | 0.000 |
| Kamonyi | 0.065 | 0.026 | 0.007 | 0.047 | 0.000 |
| Karongi | 0.063 | 0.001 | 0.000 | 0.063 | 0.000 |
| Nyamagabe | 0.060 | 0.043 | 0.000 | 0.026 | 0.000 |
| Rubavu | 0.059 | 0.028 | 0.000 | 0.031 | 0.000 |
| Gakenke | 0.058 | 0.023 | 0.036 | 0.000 | 0.000 |
| Nyaruguru | 0.055 | 0.043 | 0.000 | 0.033 | 0.000 |
| Muhanga | 0.055 | 0.019 | 0.000 | 0.036 | 0.000 |
| Nyabihu | 0.055 | 0.021 | 0.009 | 0.017 | 0.008 |
| Gisagara | 0.054 | 0.050 | 0.002 | 0.072 | 0.000 |
| Gicumbi | 0.050 | 0.019 | 0.007 | 0.024 | 0.000 |
| Kicukiro | 0.044 | 0.033 | 0.000 | 0.018 | 0.000 |
| Nyagatare | 0.042 | 0.055 | 0.000 | 0.023 | 0.000 |
| Nyanza | 0.037 | 0.036 | 0.005 | 0.016 | 0.000 |
| Nyamasheke | 0.036 | 0.036 | 0.000 | 0.000 | 0.000 |
| Rutsiro | 0.034 | 0.042 | 0.000 | 0.000 | 0.012 |
| Musanze | 0.033 | 0.017 | 0.007 | 0.009 | 0.000 |
| Bugesera | 0.032 | 0.015 | 0.001 | 0.021 | 0.006 |
| Rusizi | 0.017 | 0.008 | 0.009 | 0.000 | 0.000 |
| Rulindo | 0.015 | 0.008 | 0.000 | 0.007 | 0.000 |
| Ngororero | 0.015 | 0.007 | 0.008 | 0.008 | 0.000 |
