## Supplementary material for "Prevalence of Asymptomatic non-Falciparum and Falciparum Malaria in the 2014-15 Rwanda Demographic Health Survey": Table S6

**S6 Table. District Level Malaria Prevalence at Different Cycle Cutoffs.** Weighted n denotes the total weighted number for participants in each cluster, while all other values are district level prevalences.

| | | ***45 cycles*** | | | | | | ***40 cycles*** | | | | | |
|---|---|---|---|---|---|---|---|---|---|---|---|---|---|
| *District* | *weighted n* | *all malaria* | *non-Pf* | *Pf* | *Pm* | *Po* | *Pv* | *all malaria* | *non-Pf* | *Pf* | *Pm* | *Po* | *Pv* |
| Bugesera | 90.1 | 32.6 | 5.2 | 29.9 | 1.5 | 3.4 | 0.6 | 29.7 | 2.6 | 28.5 | 1.3 | 1.5 | 0.0 |
| Burera | 219.5 | 35.2 | 21.0 | 16.3 | 9.2 | 11.8 | 0.0 | 15.2 | 10.5 | 6.8 | 4.9 | 5.7 | 0.0 |
| Gakenke | 189.2 | 14.2 | 8.7 | 5.4 | 8.7 | 0.0 | 0.0 | 3.2 | 2.0 | 1.1 | 2.0 | 0.0 | 0.0 |
| Gasabo | 391.1 | 76.2 | 20.5 | 55.7 | 0.0 | 20.5 | 0.0 | 43.3 | 0.0 | 43.3 | 0.0 | 0.0 | 0.0 |
| Gatsibo | 110.2 | 40.0 | 15.5 | 30.4 | 0.6 | 14.9 | 0.0 | 28.4 | 4.1 | 26.0 | 0.6 | 3.5 | 0.0 |
| Gicumbi | 228.0 | 30.7 | 18.5 | 12.2 | 13.0 | 5.5 | 0.0 | 19.3 | 11.5 | 7.9 | 11.5 | 0.0 | 0.0 |
| Gisagara | 129.9 | 64.5 | 18.4 | 59.2 | 3.9 | 14.7 | 1.1 | 57.5 | 9.0 | 52.7 | 3.6 | 5.4 | 1.1 |
| Huye | 117.5 | 56.8 | 15.2 | 51.4 | 0.2 | 15.0 | 0.0 | 46.2 | 10.6 | 42.9 | 0.0 | 10.6 | 0.0 |
| Kamonyi | 137.3 | 35.2 | 10.5 | 27.9 | 1.5 | 9.0 | 0.0 | 26.2 | 3.1 | 24.3 | 0.6 | 2.6 | 0.0 |
| Karongi | 228.8 | 36.3 | 22.9 | 15.7 | 3.6 | 19.3 | 0.0 | 21.8 | 8.4 | 15.4 | 3.6 | 4.8 | 0.0 |
| Kayonza | 108.2 | 39.0 | 7.2 | 33.2 | 2.1 | 5.1 | 0.0 | 30.9 | 4.9 | 26.6 | 2.1 | 2.8 | 0.0 |
| Kicukiro | 213.9 | 29.9 | 13.3 | 17.8 | 8.9 | 5.7 | 0.0 | 20.5 | 10.8 | 10.8 | 8.9 | 1.9 | 0.0 |
| Kirehe | 102.8 | 60.6 | 13.8 | 53.7 | 5.9 | 8.4 | 0.0 | 52.1 | 9.5 | 48.4 | 5.7 | 4.0 | 0.0 |
| Muhanga | 158.0 | 26.1 | 7.3 | 20.5 | 0.9 | 6.4 | 0.0 | 17.4 | 1.7 | 17.4 | 0.9 | 0.8 | 0.0 |
| Musanze | 242.4 | 22.8 | 12.2 | 10.6 | 9.9 | 2.3 | 0.0 | 14.8 | 8.4 | 6.4 | 8.4 | 0.0 | 0.0 |
| Ngoma | 119.8 | 68.5 | 24.6 | 57.6 | 8.3 | 17.6 | 0.2 | 56.6 | 14.7 | 50.7 | 8.0 | 7.1 | 0.0 |
| Ngororero | 210.0 | 19.3 | 13.1 | 7.6 | 10.3 | 4.5 | 0.0 | 16.2 | 11.4 | 6.1 | 8.6 | 2.9 | 0.0 |
| Nyabihu | 200.1 | 21.3 | 15.4 | 6.8 | 9.4 | 4.4 | 1.6 | 10.3 | 8.6 | 2.6 | 7.6 | 1.0 | 0.0 |
| Nyagatare | 144.1 | 35.3 | 14.6 | 26.9 | 10.3 | 4.4 | 0.0 | 29.2 | 11.4 | 19.0 | 10.3 | 1.1 | 0.0 |
| Nyamagabe | 203.5 | 53.8 | 19.4 | 37.2 | 10.0 | 9.5 | 0.0 | 41.7 | 14.1 | 28.5 | 10.0 | 4.1 | 0.0 |
| Nyamasheke | 144.9 | 26.7 | 4.8 | 21.9 | 4.8 | 0.0 | 0.0 | 21.5 | 4.8 | 16.7 | 4.8 | 0.0 | 0.0 |
| Nyanza | 126.3 | 69.2 | 12.6 | 65.8 | 6.2 | 6.8 | 0.0 | 64.5 | 10.3 | 61.3 | 5.7 | 4.8 | 0.0 |
| Nyarugenge | 229.5 | 41.7 | 21.2 | 20.6 | 8.1 | 13.9 | 1.6 | 15.6 | 3.5 | 12.0 | 2.4 | 1.9 | 1.6 |

| | | | | | | | | | | | | | |
|---|---|---|---|---|---|---|---|---|---|---|---|---|---|
| Nyaruguru | 108.1 | 27.0 | 9.5 | 23.3 | 4.0 | 5.5 | 0.0 | 21.0 | 5.9 | 18.6 | 4.0 | 1.9 | 0.0 |
| Rubavu | 202.9 | 32.8 | 13.7 | 19.1 | 3.9 | 9.9 | 0.0 | 20.8 | 7.4 | 13.3 | 3.9 | 3.6 | 0.0 |
| Ruhango | 136.0 | 48.6 | 14.7 | 39.1 | 6.6 | 9.1 | 0.0 | 38.7 | 5.4 | 35.5 | 2.5 | 2.9 | 0.0 |
| Rulindo | 187.0 | 19.0 | 4.8 | 14.2 | 1.4 | 3.3 | 0.0 | 16.1 | 3.4 | 12.8 | 1.4 | 2.0 | 0.0 |
| Rusizi | 180.1 | 24.0 | 9.5 | 15.1 | 9.5 | 0.0 | 0.0 | 21.0 | 7.9 | 13.7 | 7.9 | 0.0 | 0.0 |
| Rutsiro | 132.9 | 13.8 | 5.8 | 9.1 | 4.7 | 1.1 | 1.6 | 9.2 | 5.8 | 3.4 | 4.7 | 1.1 | 0.0 |
| Rwamagana | 107.1 | 35.6 | 12.6 | 27.2 | 5.0 | 8.5 | 0.7 | 26.6 | 5.9 | 21.6 | 3.4 | 1.7 | 0.7 |
