## Supplementary material for "Prevalence of Asymptomatic non-Falciparum and Falciparum Malaria in the 2014-15 Rwanda Demographic Health Survey": Table S7

**S7 Table. Mixed Species Infection Count.** Weighted and unweighted counts at both CT value cutoffs for each category of malaria mono or co-infection among the study population.

| | |
|---|---|
| *Total unweighted count* | 5050 |
| *Total weighted count* | 5099 |

*P. falciparum infection counts*

| | ***mono infection*** | ***co-infection*** | *pf_pm* | *pf_po* | *pf_pm_po* | *pf_pm_pv* | ***total*** |
|---|---|---|---|---|---|---|---|
| *45 cycles unweighted* | 1277 | 201 | 68 | 122 | 10 | 1.0 | 1478 |
| *45 cycles weighted* | 730 | 101 | 28 | 69 | 3.5 | 1.1 | 831 |
| *under 40 cycles unweighted* | 1164 | 133 | 62 | 66 | 5.0 | 0.0 | 1297 |
| *under 40 cycles weighted* | 618 | 57 | 24 | 32 | 1.1 | 0.0 | 674 |

*P. malariae infection counts*

| | ***mono infection*** | ***co-infection*** | *pf_pm* | *pm_po* | *pm_pv* | *pf_pm_po* | *pf_pm_pv* | ***total*** |
|---|---|---|---|---|---|---|---|---|
| *45 cycles unweighted* | 99 | 74 | 56 | 5.0 | 3.0 | 9.0 | 1.0 | 173 |
| *45 cycles weighted* | 131 | 41 | 28 | 5.0 | 4.0 | 3.5 | 1.1 | 172 |
| *under 40 cycles unweighted* | 89 | 70 | 62 | 1.0 | 2.0 | 5.0 | 0.0 | 159 |
| *under 40 cycles weighted* | 111 | 28 | 24 | 0.8 | 2.8 | 1.1 | 0.0 | 139 |

*P. ovale infection counts*

| | ***mono infection*** | ***co-infection*** | *pf_po* | *pm_po* | *pf_pm_po* | ***total*** |
|---|---|---|---|---|---|---|
| *45 cycles unweighted* | 146 | 137 | 122 | 5.0 | 9.0 | 283 |

| | | | | | | |
|---|---|---|---|---|---|---|
| *45 cycles weighted* | 163 | 77 | 69 | 5.0 | 3.5 | 241 |
| *under 40 cycles unweighted* | 52 | 72 | 66 | 1.0 | 5.0 | 124 |
| *under 40 cycles weighted* | 46 | 34 | 32 | 0.8 | 1.1 | 80 |

*P. vivax infection counts*

| | ***mono infection*** | ***co-infection*** | *pm_pv* | *pf_pm_pv* | ***total*** |
|---|---|---|---|---|---|
| *45 cycles unweighted* | 3.0 | 4.0 | 3.0 | 1.0 | 7.0 |
| *45 cycles weighted* | 2.4 | 5.1 | 4.0 | 1.1 | 7.5 |
| *under 40 cycles unweighted* | 1.0 | 2.0 | 2.0 | 0.0 | 3.0 |
| *under 40 cycles weighted* | 0.7 | 2.8 | 2.8 | 0.0 | 3.5 |

*All malaria infection counts*

| | ***mono infection*** | ***co-infection*** | *total pf* | *total pm* | *total po* | *total pv* | *total non-pf* | ***total*** |
|---|---|---|---|---|---|---|---|---|
| *unweighted count* | 1525 | 389 | 1478 | 173 | 283 | 7 | 463 | 1941 |
| *weighted count* | 1026 | 225 | 831 | 172 | 241 | 7.5 | 420 | 1251 |
| *under 40 CT unweighted count* | 1306 | 277 | 1297 | 159 | 124 | 3 | 286 | 1583 |
| *weighted under 40 count* | 775 | 121 | 674 | 139 | 80 | 3.5 | 222 | 897 |
